# Executive Function as an Underlying Mechanism of Alcohol Use, Aggression, and ADHD

**DOI:** 10.1101/2024.06.10.24308620

**Authors:** Kellyn M. Spychala, Naomi P. Friedman, Ian R. Gizer

**Author notes:** Correspondence concerning this article should be addressed to Kellyn Spychala, Department of Psychological Sciences, University of Missouri, Columbia, MO 65211.

## Abstract

**Background:** Executive functioning (EF) has been proposed as a transdiagnostic risk factor for externalizing disorders and behavior more broadly, including attention-deficit/hyperactivity disorder (ADHD), aggression, and alcohol use. Previous research has demonstrated both phenotypic and genetic overlap among these behaviors, but has yet to examine EF as a common causal mechanism. The current study examined reciprocal causal associations between EF and several externalizing behaviors using a Mendelian randomization (MR) approach.

**Methods:** Two-sample MR was conducted to test causal associations between EF and externalizing behaviors. Summary statistics from several genome-wide association studies (GWASs) were used in these analyses, including GWASs of EF, ADHD diagnostic status, drinks per week, aggressive behavior, and alcohol use disorder (AUD) diagnostic status. Multiple estimation methods were employed to account for horizontal pleiotropy (e.g., inverse variance weighted, MR-PRESSO, MR-MIX).

**Results:** EF demonstrated significant causal relationships with ADHD (P < 0.01), AUD (P < 0.03), and alcohol consumption (P < 0.01) across several estimation methods. Reciprocally, ADHD showed a significant causal influence on EF (P < 0.03). Nonetheless, caution should be used when interpreting these findings as there was some evidence for horizontal pleiotropy in the effect of EF on ADHD and significant heterogeneity in variant effects in the other relations tested. There were no significant findings for aggression.

**Conclusions:** Findings suggest that EF may be a causal mechanism underlying some externalizing behaviors, including ADHD and alcohol use, and that ADHD may also lead to lower performance on EF tasks.

## Introduction

Many psychiatric disorders, such as substance use disorders, attention-deficit/hyperactivity disorder (ADHD), and conduct disorder (CD)/antisocial personality disorder, along with some personality traits, including trait aggression, have been shown to co-occur frequently throughout the lifespan (Krueger et al., 2007). Individual behaviors and traits associated with these disorders in childhood and adolescence (e.g., impulsive action, aggressive behavior, substance use), as well as their co-occurrence, predict greater rates of criminal offenses among adults (Magee et al., 2021; Moore et al., 2019). Additionally, these co-occurring behaviors are associated with increased rates of hospitalizations among adolescents (Masroor et al., 2019) and increased rates of suicide attempts independent of comorbid internalizing psychopathology (Verona et al., 2004; Commisso et al., 2023). Together, these outcomes result in significant public costs, with an estimated additional $70,000 in public expenditures made for a child diagnosed with CD compared to a child without CD during adolescence in the US (Foster et al., 2005). Similarly, a study conducted in Finland reported that, over a 23-year period, roughly 4 times as much is spent on a child with a greater level of conduct problems (€44,348/$47,600) compared to a child with low levels of these problems (€10,547/$11,320; Rissanen et al., 2022). Given the associated suffering and public health costs, it is imperative to understand the underlying mechanisms of these behaviors, including mechanisms that may explain their co-occurrence.

Latent variable models capture the covariance among these behaviors with a factor commonly referred to as the externalizing spectrum (Achenbaeh TM and Edelbrock C, 1991; Krueger et al., 2005). While evidence suggests that aggressive/antisocial and substance use behaviors may represent separable subfactors within the externalizing spectrum, there is also substantial evidence indicating these facets are highly correlated and can be subsumed under a single factor (Ingole et al., 2015; Krueger et al., 2007; Poore et al., 2023). Together, such findings suggest that these behaviors have shared underlying mechanisms, but also raise the possibility of heterogeneity in the relations between these mechanisms and individual externalizing behaviors.

Executive functioning (EF) deficits represent one proposed shared mechanism of externalizing behavior (Beauchaine et al., 2014). For example, a metanalysis of longitudinal studies demonstrated that early executive dysfunction was associated with greater ADHD and CD symptoms and increased substance use in later childhood/adolescence (Yang et al., 2022). Further, another metanalysis demonstrated that interventions targeting specific EF processes (i.e., inhibitory control, working memory, flexibility) were able to reduce symptoms of externalizing behaviors (i.e., ADHD, oppositional defiant disorder) in children (Pauli-Pott et al., 2021). Together, these studies suggest that EF plays a critical role in the manifestation and maintenance of several externalizing behaviors and associated disorders.

As alluded to in the prior paragraph, EF is frequently described as a multifaceted construct representing a collection of processes that allow for goal directed behavior (Suchy, 2009). Three separable and commonly studied EF processes are inhibition, working memory, and set-shifting or cognitive flexibility (Diamond, 2013; Friedman & Miyake, 2017; Miyake et al., 2000). In terms of their neural correlates, these processes have been most prominently localized to the prefrontal cortex (PFC) and the anterior cingulate cortex (ACC), both parts of the frontal lobe. The dorsolateral PFC in particular plays a critical role in these processes with localized activation of this area and connectivity between this area and the ACC increasing as EF develops across the lifespan (Fiske & Holmboe, 2019).

These findings have motivated studies exploring whether disruptions in the PFC and ACC are associated with externalizing behaviors (Beauchaine et al., 2015). For, example, a metanalysis demonstrated that ADHD, which is characterized by impairments in EF (Lawrence et al., 2004), was associated with decreases in PFC and ACC activation during attention and inhibition tasks, respectively (Hart et al., 2013). Reduced activation of these areas is also associated with aggressive behavior resulting from emotional arousal, and alcohol consumption can reduce activation of the PFC, resulting in an increased tendency towards aggression (Heinz et al., 2011). There is also a long-term effect of chronic alcohol use on EF and PFC functioning, suggesting a bidirectional relationship between alcohol use and EF (Heinz et al., 2011).

In addition to functional differences, structural neuroimaging studies have suggested relations between PFC and ACC and EF and externalizing behavior. One such study found reduced grey matter volume in the ACC among healthy controls with deficits in EF and among individuals with psychopathology, including substance use disorders (Goodkind et al., 2015). Another study reported that associations between externalizing behavior (i.e., ADHD and conduct problems) and white matter microstructure were mediated by EF (Cardenas-Iniguez et al., 2022), suggesting that EF deficits may explain the link between structural differences in brain and externalizing behavior. Thus, both functional and structural neuroimaging studies provide robust evidence of shared biological functions related to EF underlying externalizing behavior.

Genetic approaches can also be used to investigate the underpinnings of externalizing behaviors, including the influence of EF. Twin studies demonstrate that both normative alcohol consumption, problematic alcohol use (e.g., AUD), and aggression show modest heritability estimates among adults (around 50%; Hansell et al., 2008; Hudziak et al., 2000, 2003; Rose & Dick, 2005; Tellegen et al., 1988; Verhulst et al., 2015). In addition, twin studies have demonstrated that ADHD is influenced by genetic factors, with heritability estimates between 70 and 80% (Faraone & Larsson, 2019). While individual EF tasks have a wider range of heritability estimates (29-76%), the heritability of latent EF factors modeling the covariance among tasks is uniform and high (81-96%; Friedman et al., 2016). Importantly, twin studies have also demonstrated moderate genetic correlations between alcohol dependence and aggressive behavior (McAdams et al., 2012; von der Pahlen et al., 2008), ADHD (*r_g_* = 0.16-0.50; Derks et al., 2014; Quinn et al., 2016), and CD (Slutske et al., 1998), with the latter also showing moderate genetic correlations with EF (*r_g_* = 0.51; Coolidge et al., 2004) and ADHD (*r_g_* = 0.17-0.70; Burt et al., 2001; Silberg et al., 2015), providing further evidence of shared etiology among EF and externalizing behavior.

More recently, molecular genetics approaches, such as linkage disequilibrium score regression (LDSC: Bulik-Sullivan et al., 2015), have demonstrated genetic overlap between EF, ADHD, aggression, and alcohol use (Marees et al., 2020). For example, ADHD is positively genetically correlated with aggression (Manchia & Fanos, 2017; Retz & Rösler, 2009) and alcohol use quantity (Marees et al., 2020), though negatively correlated with alcohol use frequency (Marees et al., 2020). On the other hand, cognitive functioning is negatively genetically correlated with both alcohol use frequency and quantity (Marees et al., 2020). Finally, one study using genomic structural equation modelling (Grotzinger et al., 2019), which extends the LDSC approach to allow for the modeling of latent genetic variables, demonstrated associations between an addiction factor, defined by substance use disorders, and EF and early neuro-developmental disorders, including ADHD, further suggesting shared genetic underpinnings among these constructs (Hatoum et al., 2022).

Given the described evidence, it is suspected that causal relationships exist between EF and externalizing behavior (Snyder et al., 2015). As described above, the Yang et al. (2022) metanalysis of longitudinal studies provides evidence for temporal precedence in which EF deficits predict subsequent externalizing behavior. However, other longitudinal studies have found reverse relationships in which early externalizing behavior predicted later deficits in EF but early EF did not predict later externalizing behavior (Brieant et al., 2022; Dontati et al., 2021). Conflicting findings among longitudinal studies suggest potential transactional relations between EF and externalizing behavior, motivating the examination of the reciprocal nature of these relationships using causal modeling approaches.

Mendelian randomization (MR), a form of instrumental variable analysis, can be used to test for causal relationships between an exposure and an outcome, as long as the exposure is significantly influenced by genetic variants that can then be used as the instrumental variables (Sanderson et al., 2022). As genetic variants are randomly assorted to offspring from each parent, mimicking random assignment in a randomized control trial, it follows that a variant’s association with the proposed exposure and outcome provides a test of causality under the key assumption that the genetic variant is associated with the outcome solely through the variant’s effect on the exposure. If the genetic variant exhibits direct effects on both the exposure and outcome (i.e., horizontal pleiotropy), this assumption is violated and inferences regarding causality cannot be made.

The MR approach has provided some isolated evidence for causal relationships between externalizing behaviors, such as alcohol use playing a causal role in aggression (Chao et al., 2017), and that ADHD may play a causal role in alcohol use (Treur et al., 2021). However, there has been limited research on the effects of EF on externalizing behavior using causal modeling methods. One MR study demonstrated a causal relationship between EF and alcohol consumption but did not find significant evidence for a causal relationship between EF and AUD (Burton et al., 2022). However, this study did not examine relations between EF and non-substance use related externalizing behaviors.

Thus, the current study sought to extend this work by examining whether there were reciprocal causal relations between EF and several externalizing behaviors, including alcohol use, AUD, aggression, and ADHD using MR methods. Given that impairments in EF are a key characteristic of ADHD (Lawrence et al., 2004) and EF impairments are associated with greater alcohol use (Day et al., 2015) and aggression (Poland et al., 2016), it was hypothesized that EF would show negative causal relationships with these phenotypes. Given the limited evidence for externalizing behaviors causing decreases in EF, those analyses were considered exploratory.

## Methods

### Data

MR analyses were conducted using sets of GWAS summary statistics for aggression (n = 87,485; Ip et al., 2021), alcohol consumption (n = 403,931; obtained from direct-to-consumer genetics company 23andMe, Inc., Sunnyvale, CA and described in Liu et al., 2019), AUD (n = 514,944), ADHD (n= 225,534; Demontis et al., 2023), and EF (n = 427,037; Hatoum et al., 2023). For the AUD summary statistics, a metanalysis of separate summary statistics from the Psychiatric Genomics Consortium (Walters et al., 2018), Million Veterans Program (Zhou et al., 2020), and FinnGen (FinnGen, 2021) was conducted (Spychala et al., 2024). These GWAS summary statistics were chosen in order to avoid sample overlap, which can bias MR analyses. See Table 1 for sample information from summary statistics used in this study.

**Table 1.** Descriptive Information for GWAS Summary Statistics.

| **Phenotype** | **Authors** | **Samples** | **N** |
| --- | --- | --- | --- |
| Executive Function | Hatoum et al. (2023) | UKBiobank | 427,037 |
| Alcohol Consumption | Liu et al. (2019) | 23andMe | 403,931 |
| Alcohol Use Disorder | Spychala et al. (2024) | Psychiatric Genetics Consortium, Million Veterans Program, FinnGen | 514,944 |
| Aggression | Ip et al. (2021) | 29 cohorts (see article for full descriptions of samples) | 87,485 |
| Attention-Deficit/Hyperactivity Disorder | Demontis et al. (2023) | iPSYCH, deCODE, Psychiatric Genetics Consortium | 225,534 |

### Statistical Analyses

Two-sample MR analyses were primarily implemented in the TwoSampleMR R package (Hemani, Zheng, et al., 2018) with additional packages described below. SNPs with p-values < 5 x 10^-8^ (or < 1 x 10^-5^ in the case of the aggression summary statistics, which do not have genome-wide significant SNPs) were selected as exposures. Using this approach, causality was tested between alcohol consumption, AUD, aggression, ADHD, and EF phenotypes in both directions. As an example, in models that test the EF-to-aggression relation, SNPs were extracted based on significance in the EF discovery GWAS (instruments measuring exposure). Effects for these SNPs were then extracted from the aggression target GWAS (instruments measuring outcome). It is assumed that if the selected variants influence aggression only indirectly through their effect on EF, their effects on the aggression phenotype should be proportional to their effects on EF (Sanderson et al., 2022). Parallel models were used to test the aggression-to-EF relation and reciprocal relations between EF and all other externalizing phenotypes.

Given that the “no horizontal pleiotropy” assumption of MR can be difficult to evaluate, multiple MR methods were used to conduct these analyses. Specifically, five estimation methods available in TwoSampleMR – inverse variance weighted (IVW), weighted mode, simple mode, weighted median, and MR Egger regression methods (Jones et al., 2020), each with different assumptions regarding the validity of the instruments, were employed to analyze each MR model. We also used methods that are robust to variants that act as outliers in the analysis (e.g., MR-PRESSO; Verbanck et al., 2018) and more recently developed methods that attempt to model the distributions of variants as valid and invalid instruments (e.g., MR-Mix R package; Qi & Chatterjee, 2019). Each approach has specific strengths and weaknesses (Slob & Burgess, 2020).

Sensitivity analyses were also conducted to test whether the assumptions of MR were upheld. These included a test for heterogeneity between SNP effects using Cochran’s Q and examination of the intercept from MR-Egger to test for horizontal pleiotropy (Hemani et al., 2017).

## Results

### Causal effects of EF on alcohol use, aggression, and ADHD

There was some evidence of decreased EF leading to a higher likelihood of an AUD diagnosis (see Table 2 and Figure 1 for full results). This was demonstrated across several estimation methods: weighted median (b= −0.14, P= 0.03, CI: −0.004to −0.27), IVW (b= −0.13, P= 0.03, CI: −0.006 to −0.25), and the MR-PRESSO estimate after outlier correction (b= - 0.12, P= 0.02). The MR-PRESSO distortion test also suggested that causal estimates were not significantly influenced by outliers (P= 0.88). Sensitivity analyses did not suggest any significant horizontal pleiotropy (MR-Egger - P = 0.19), but did suggest some heterogeneity in SNP effects (Cochrane’s Qs - P < 8.3 x 10^-9^).

**Figure 1.**
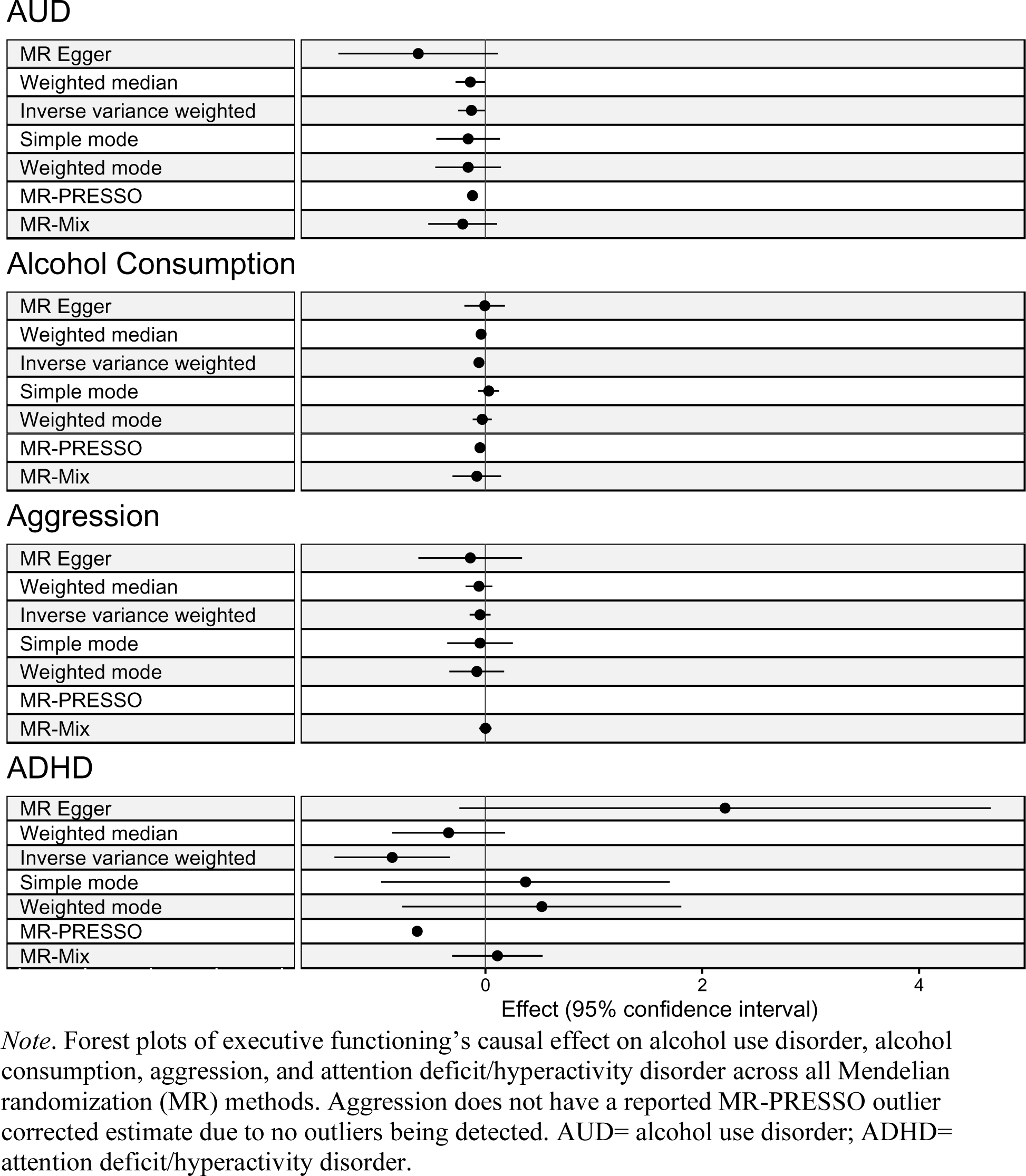
Executive Functioning’s Causal Effects on Externalizing Phenotypes across Mendelian Randomization Methods.

**Table 2.** Causal Effects of Executive Functioning on Alcohol Use, Aggression, and ADHD.

| Method | AUD |  |  | Alcohol Consumption |  |  | Aggression |  |  | ADHD |  |  |
| --- | --- | --- | --- | --- | --- | --- | --- | --- | --- | --- | --- | --- |
| | $\beta$ | SE/SD | *P* | $\beta$ | SE/SD | *P* | $\beta$ | SE/SD | *P* | $\beta$ | SE/SD | *P* |
| MR Egger | -0.62 | 0.376 | 0.11 | -0.007 | 0.095 | 0.93 | -0.14 | 0.244 | 0.55 | 2.21 | 1.25 | 0.08 |
| Weighted Median | -0.14 | 0.069 | **0.03** | -0.04 | 0.019 | **0.01** | -0.06 | 0.063 | 0.31 | -0.34 | 0.266 | 0.19 |
| IVW | -0.13 | 0.063 | **0.03** | -0.06 | 0.020 | **0.001** | -0.05 | 0.049 | 0.31 | -0.86 | 0.272 | **0.001** |
| Simple Mode | -0.16 | 0.149 | 0.28 | -0.03 | 0.049 | 0.51 | -0.05 | 0.154 | 0.73 | 0.37 | 0.679 | 0.59 |
| Weighted Mode | -0.16 | 0.154 | 0.30 | -0.03 | 0.045 | 0.47 | -0.08 | 0.129 | 0.49 | 0.52 | 0.656 | 0.42 |
| MR-PRESSO after outlier correction<sup>+</sup> | -0.12 | 0.056 | **0.02** | -0.05 | 0.017 | **0.001** | * |  |  | -0.63 | 0.240 | **0.01** |
| MR-Mix | -0.21 | 0.162 | 0.19 | -0.08 | 0.114 | 0.48 | 2x10<sup>-17</sup> | 0.029 | 0.99 | 0.11 | 0.213 | 0.61 |

Note: Mendelian randomization (MR) analyses were conducted across 7 methods. Significant results are denoted by bold text. \* - indicates there were no significant outliers to conduct the MR-PRESSO outlier correction test. + - all methods report standard errors with the exception of MR-PRESSO which reports the standard deviation. Abbreviations: AUD= alcohol use disorder, ADHD= attention-deficit/hyperactive disorder, IVW= inverse variance weighted, SE = standard error, SD = standard deviation.

Similarly, several estimation methods suggested evidence of decreased EF leading to greater alcohol consumption: weighted median (b= −0.04, P= 0.017, CI: −0.002 to −0.07), IVW (b= −0.06, P= 0.001, CI: −0.02 to −0.09), and the MR-PRESSO estimate after outlier correction (b= −0.05, P= 0.001). The MR-PRESSO distortion test also suggested that causal estimates were not significantly influenced by outliers (P= 0.71). Sensitivity analyses did not suggest any significant horizontal pleiotropy (MR-Egger - P = 0.55), but did suggest some heterogeneity in SNP effects (Cochrane’s Qs - P < 1.9 x 10^-17^).

Finally, there was weak evidence of decreased EF leading to a higher likelihood of an ADHD diagnosis (see Table 2 for full results) using the IVW method (b= −0.86, P= 0.001, CI: −0.32to −1.39), and the MR-PRESSO estimate after outlier correction (b= −0.63, P= 0.01). The MR-PRESSO distortion test also suggested that causal estimates were not significantly influenced by outliers (P= 0.16). However, sensitivity analyses suggested the presence of horizontal pleiotropy (MR-Egger - P = 0.01) and heterogeneity in SNP effects (Cochrane’s Qs - P < 9.4 x 10^-14^). There was no evidence for a causal effect of EF on aggression (see Table 2 for full results).

### Causal effects of alcohol use, aggression, and ADHD on EF

There was evidence of an ADHD diagnosis leading to decreased EF task performance across several estimation methods: weighted median (b= −0.03, P= 0.000001, CI: −0.04 to −0.01), IVW (b= −0.03, P= 0.00009, CI: −0.04 to −0.01), simple mode (b= −0.04, P= 0.01, CI: −0.06 to −0.01), weighted mode (b= −0.03, P= 0.03, CI: −0.05 to −0.001), the MR-PRESSO estimate after outlier correction (b= −0.03, P= 0.0007), and MR-Mix (b= −0.09, P= 0.000001, CI: −-0.12 to −0.05). Sensitivity analyses indicated heterogeneity in SNP effects (Cochrane’s Qs - P < 3.97 x 10^-7^), but no horizontal pleiotropy (MR-Egger - P=0.77). The MR-PRESSO sensitivity analyses also indicated that there was no significant change in the estimated effect after the removal of outliers (see Table 3 for full results).

**Table 3.**
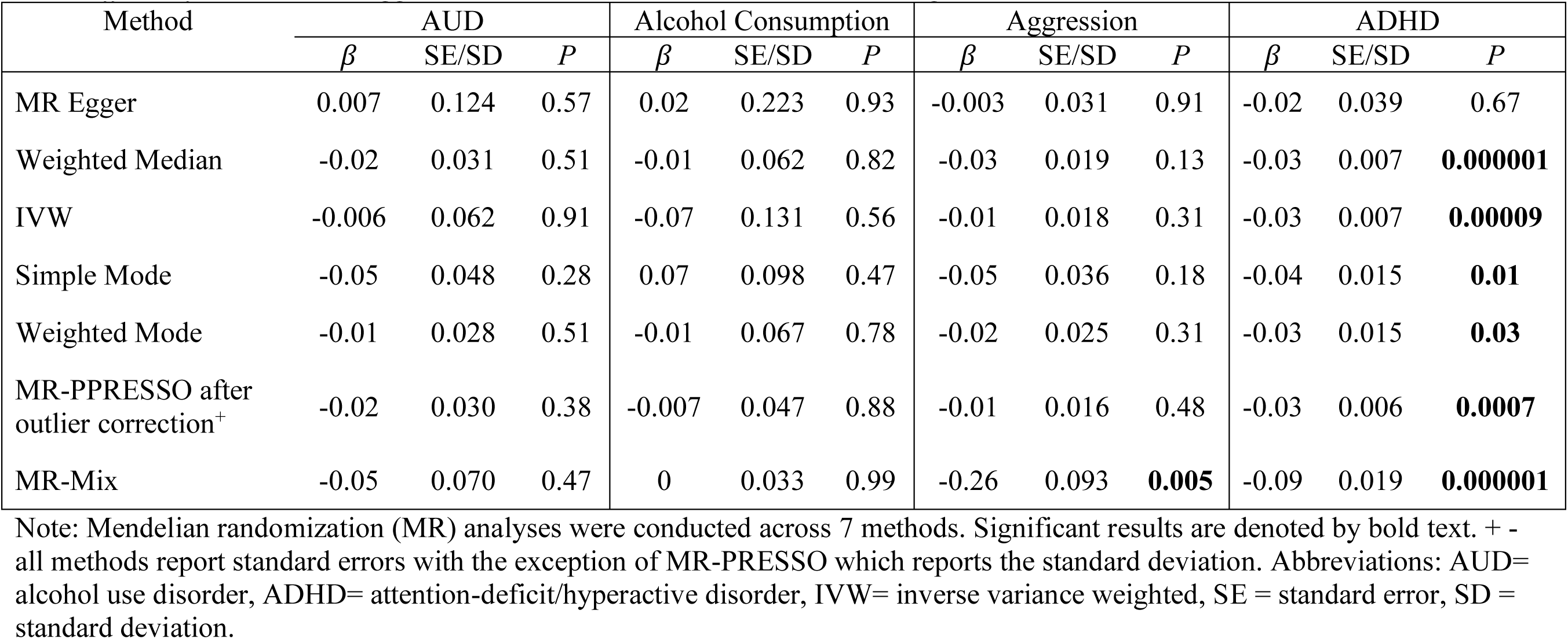
Causal Effects of Alcohol Use, Aggression, and ADHD on Executive Functioning.

Additionally, there was evidence for a causal effect of aggression on EF using the MR- Mix approach (b= −0.26, P= 0.005, CI: −0.44 to −0.07), though this evidence was viewed as weak given that it was the only significant finding across MR methods. There was no evidence supporting a causal effect of alcohol consumption or AUD on EF (see Table 3 for full results).

## Discussion

The present study examined the causal relations between EF and several externalizing behaviors using an MR approach to test the hypothesis that EF represents a shared underlying causal mechanism in the etiology of these behaviors. Multiple MR estimation methods were employed to evaluate consistency in results across methods that differently account for the instrumental variable assumptions, thus allowing for greater confidence when a pattern of significant results indicated the presence of a causal relationship. While there is one previous MR study examining the reciprocal relations between EF and psychological disorders, including alcohol use and AUD (Burton et al., 2022), the present study provided several important extensions of this earlier work. First, the present study included a broader range of externalizing behaviors (i.e., ADHD and aggression) when examining relations with EF. Second, the summary statistics used for the MR analyses in the present study included larger samples relative to those used in the prior study (i.e., AUD). Third, a wider range of MR methods were employed (i.e., MR-PRESSO and MR-Mix) to evaluate the robustness of observed relations.

Given the strong evidence for EF deficits predicting several externalizing behaviors (Pinsonneault et al., 2015), the causal influences of EF on aggression, alcohol consumption, AUD, and ADHD, were examined first. Across several estimation methods, there was evidence for EF causally influencing AUD (P < 0.03), alcohol consumption (P <0.01), and ADHD (P < 0.01). These results provide support for prior behavior genetic and neuroimaging studies (Cardenas-Iniguez et al., 2022; Coolidge et al., 2004; Goodkind et al., 2015; Hatoum et al., 2022; Marees et al., 2020; Silberg et al., 2015), suggesting that behaviors on the externalizing spectrum share common underlying causal mechanisms and that EF may represent one such mechanism. This finding is consistent with previous longitudinal studies providing evidence that early deficits in EF predict later substance use and the onset of ADHD symptoms (Yang et al., 2022). However, these findings are contrary to the one MR study in which EF did not demonstrate significant causal associations with AUD (Burton et al., 2022). These discrepant results can likely be explained, at least in part, by the use of different GWAS summary statistics for AUD between the two studies, with summary statistics used in the present study derived from a larger overall sample. Together, the findings from the present study suggest that intervening on EF impairments may reduce the risk of developing several externalizing disorders, including alcohol consumption, AUD, and ADHD, consistent with the limited intervention research in this area (Pauli-Pott et al., 2021).

The second set of findings to emerge from the present study was the presence of causal effects of externalizing behavior on EF task performance. Across several estimation methods, ADHD demonstrated significant causal influences on EF, which is inconsistent with theories that early deficits in EF predict later development of externalizing behavior (Hughes and Ensor, 2008). However, there has been some evidence from longitudinal studies that the relationship between EF and externalizing behavior may be transactional, in that EF may lead to the development of externalizing behavior which in turn predicts lower EF later in development (Brieant et al., 2022; Donati et al., 2021). Evidence that psychopathology can have negative causal influences on cognitive function have also been observed outside of the externalizing spectrum, with one study suggesting such an effect of schizophrenia on intelligence (Ohi et al., 2021). These studies suggest potentially reciprocal and transactional relations between cognitive function and psychopathology, but the precise mechanisms of these relations cannot be teased apart solely using MR methods. For example, long-term effects of ADHD could include the development of later neurodegenerative diseases associated with greater EF dysfunction (Becker et al., 2022), or it is possible that some types of task performance (i.e., attention tasks) can be influenced by decreased levels of motivation among those diagnosed with ADHD compared to healthy controls (Dekkers et al., 2017). Unfortunately, due to limitations of the MR approach, such nuance cannot be teased apart, thus demonstrating the need for longitudinal studies that can further disentangle the causal relationships between ADHD and EF and test for transactional relationships between EF and externalizing behavior.

It should be noted that there are limitations of the current study that need to be considered when interpreting the findings. There was evidence of horizontal pleiotropy in the models examining EF’s causal influence on ADHD, which suggests that this relationship may not be causal but rather that some of the variants are invalid instruments influencing both EF and ADHD directly, a plausible alternative hypothesis to a causal relationship between EF and ADHD. The evidence for horizontal pleiotropy may also explain the demonstrated variability across the different MR methods in the direction of the effect estimates (i.e., negative and positive effects) when examining the causal influence of EF on ADHD. A similar limitation is the evidence for heterogeneity among variant effects in many of the examined models. While this heterogeneity could suggest a violation of the horizontal pleiotropy assumption of MR among one or all instrumental SNPs, heterogeneity can also occur for other reasons, including the heterogeneity tests being biased towards false positives (Hemani, Bowden, et al., 2018). The lack of significant results from the MR-PRESSO distortion tests provides evidence that the heterogeneity may be a result of the latter rather than horizontal pleiotropy.

An additional limitation of this study is the smaller sample size of the GWAS summary statistics for aggression, which may have contributed to the lack of significant findings for this phenotype. The aggression GWAS only included 87,485 individuals, much fewer than the other traits. Due to its smaller sample size, this GWAS also did not detect any SNPs at the genome-wide significance level, and thus, a threshold of P < 1 x 10^-5^ had to be used for the present study. Using a more liberal threshold potentially results in weaker instruments for the aggression phenotype and thus, limits the ability to detect causal effects on EF (Burgess and Thompson, 2011).

Finally, all of the GWAS summary statistics used in this study were derived from individuals of European ancestry. Exclusion of individuals of other ancestries, was necessary because some of these GWASs (i.e., aggression and EF) have only been conducted in individuals of European ancestry and MR methods require that both sets of summary statistics be derived from individuals of the same ancestry group to avoid bias that can result from different patterns in linkage disequilibrium across ancestral populations (Hemani, Zheng, et al., 2018). Nonetheless, it is possible that different causal relationships could emerge across ancestral groups, and thus future research should focus on conducting GWAS of these traits in multi-ancestry samples.

Despite these limitations, the current study provides evidence of EF having causal relationships on alcohol consumption and AUD, as well as a potentially bidirectional causal relationship with ADHD. Future research should focus on evaluating whether individual genomic segments provide evidence of causal relations between these traits to identify biological mechanisms underlying EF that may give rise to the observed causal relations with AUD and alcohol consumption. Notably, identifying those genetic variants with pleiotropic effects would also help to further our understanding of the biological relationships between EF and ADHD. Additionally, longitudinal studies with more assessments across the lifespan are warranted to more fully elucidate the timing of changes in EF and the onset of ADHD and AUD symptoms. Studies that examine genetic effects on these phenotypes at different ages could also contribute to future MR studies and provide additional information on the directionality and timing of causal effects.

## Data Availability

The full GWAS summary statistics for the 23andMe discovery data set will be made available through 23andMe to qualified researchers under an agreement with 23andMe that protects the privacy of the 23andMe participants. Please visit https://research.23andme.com/collaborate/#dataset-access/ for more information and to apply to access the data.
PGC alcohol dependence GWAS summary statistics were obtained from the PGC website (https://www.med.unc.edu/pgc/). Million Veteran Program GWAS summary statistics were obtained through the Database for Genotypes and Phenotypes (dbGaP; Study Accession: phs001672). FinnGenR5 ICD-based AUD GWAS data were obtained from https://www.finngen.fi/en/access_results.
The executive functioning GWAS summary statistics are available at http://ftp.ebi.ac.uk/pub/databases/gwas/summary_statistics/GCST90162001-GCST90163000/GCST90162547. The aggression GWAS summary statistics are available at https://tweelingenregister.vu.nl/eagle-gwa-meta-analyses-summary-results.

https://www.finngen.fi/en/access_results

http://ftp.ebi.ac.uk/pub/databases/gwas/summary_statistics/GCST90162001-GCST90163000/GCST90162547

https://tweelingenregister.vu.nl/eagle-gwa-meta-analyses-summary-results

## Acknowledgements

This study made use of summary statistics data from a number of sources. First, this study used GWAS summary statistics data from 23andMe, Inc. (Sunnyvale, CA). We thank the 23andMe research participants and employees for making this work possible. Second, this research used summary data from the UK Biobank, a population-based sample of participants whose contributions we gratefully acknowledge. Third, this study used GWAS summary statistics data from the fifth release of the FinnGen study. We thank the participants and investigators of the FinnGen study. Fourth, this research also used summary data from the Psychiatric Genomics Consortium Substance Use Disorders (PGC-SUD) working group. PGC– SUD is supported by funds from NIDA and NIMH to MH109532 and, previously, had analyst support from NIAAA to U01AA008401 (COGA). PGC–SUD gratefully acknowledges its contributing studies and the participants in those studies, without whom this effort would not be possible. Fifth, this research used data from the Million Veteran Program and was supported by funding from the Department of Veterans Affairs Office of Research and Development, Million Veteran Program Grant nos. I01BX003341 and I01CX001849; and the VA Cooperative Studies Program study, no. 575B. This publication does not represent the views of the Department of Veterans Affairs or the United States Government. Finally, this study used summary statistics data from the ACTION consortium. We gratefully acknowledge the ACTION participants for their contributions.

